# Reaching the Remote: Dried blood spot analysis for disease diagnosis on a protein microarray platform

**DOI:** 10.1101/2022.05.09.22274830

**Authors:** Metoboroghene O. Mowoe, Tristan Rensburg, Hisham Ali, Joshua Gqada, Urda Kotze, Marc Bernon, Bradley Africa, Eduard Jonas, Jonathan M. Blackburn

**Affiliations:** Department of Integrative Biomedical Sciences, Division of Chemical and Systems Biology; Institute of Infectious Disease & Molecular Medicine, University of Cape Town, Cape Town, South Africa; Surgical gastroenterology Unit, Division of General Surgery, Groote Schuur Hospital, University of Cape Town, Cape Town, South Africa

## Abstract

Cancer remains one of the leading causes of death globally with an estimated 19.3 million cases and 10 million mortalities in 2020. In Africa and Asia, where remoteness is prevalent, access to healthcare facilities is limited, providing a significant barrier to effective screening and early detection of cancers in at-risk groups and thus, incomplete registries. Here, we utilised low resource, low-cost dried blood spots (DBS)-based sample collection coupled with robust, protein microarray technology to enable quantitative, multiplexed measurements of diagnostic autoantibody biomarkers of disease, in minimal sample volumes. Specifically, we describe the development of a DBS extraction and elution method from low cost, home-made blood cards. We then show that DBS stored at room temperature (25 □, RT) for up to 15 d yield comparable autoantibody signatures to autologous serum samples stored at -80 □ and those from samples prepared on a commercially available blood card. We further conducted a pilot study, comparing total IgG and three previously identified autoantibodies upregulated in pancreatic cancer (PC), in DBS from 11 PC patients stored at RT for up to 15 d. We found comparable protein profiles across commercially developed blood cards and our low cost, in-house kit with no significant difference in autoantibody profiles over 15 d (p > 0.05). Such low cost, DBS-based sample collection methods, combined with regular, RT courier shipments and ultrasensitive protein microarray-based detection in a remote laboratory, thus have the potential to facilitate future, unbiased, large scale serosurveys and serological diagnostic testing within remote, rural communities.

## INTRODUCTION

Geographic variations in cancer incidence, and consequentially survival, are partly due inequity and inequality in terms of global access to healthcare and the quality of registries in remote, rural populations ^1,2^. Thus, reported disease incidence rates are often inversely correlated to rural population proportions in different regions (Figure 1), despite a worse health status in remote, rural areas than urban areas ^3,4^. This implies dramatic under-reporting in such areas due to limited access to screening and is most prominent in middle- and low-income countries (LMICs), most of which exist on the African and Asian continents.

**Figure 1.**
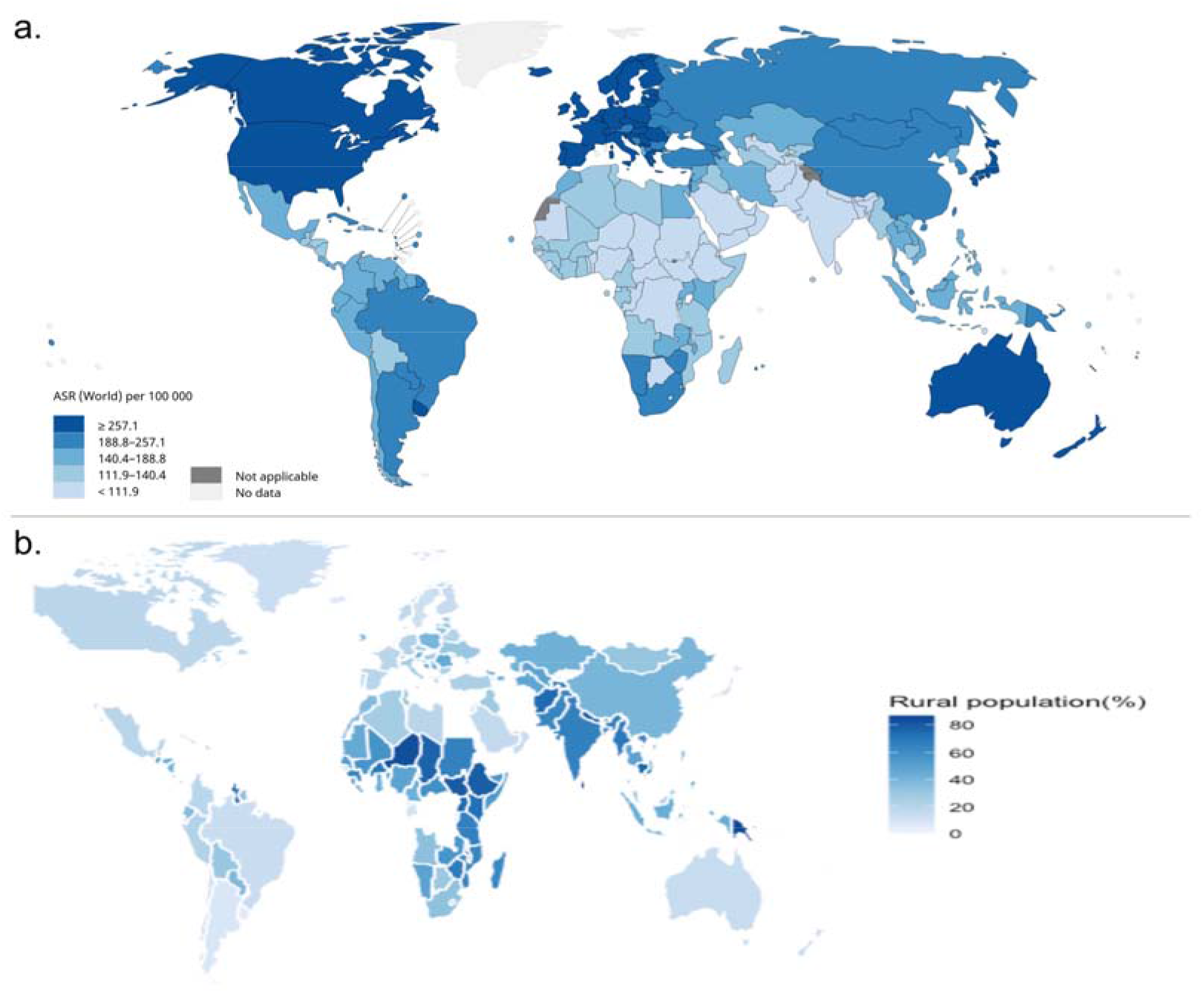
Maps of global cancer incidence and rural population percentage a.) Estimated age-standardized global incidence rates for all cancers, including both sexes and ages in 2020 (produced in http://gco.iarc.fr/today/home) ^9^, b.) Percentage of global rural population (data derived from https://data.worldbank.org/indicator/SP.RUR.TOTL.ZS?end=2019&start=1960&view=map) and plotted using ggplot ^10^ in R.

Limited access to diagnostic tools, funding for diagnostic and laboratory services and specialists, combined with patient-related barriers to follow through with diagnostic referrals, impede disease diagnosis in these communities ^5^. To overcome diagnostic limitations, dried blood spots (DBS) have been used as an analytical matrix in the clinical setting for over half a century, primarily for disease screening of neonates ^6-8^. They are minimally invasive, require little to no training and storage infrastructure, and obviate the risk associated with needles and syringes used in venous blood collection. However, the minimal recovered sample volumes can create technical challenges for downstream protein biomarker measurements that lack the massive signal amplification of PCR methods.

In cancers, autoantibodies (Aabs) have gained recent popularity as candidate biomarkers since they exhibit increased levels during the early stages of disease and can potentially predict disease progression and treatment outcomes^11^. Increases in Aab concentration may be detectable months to years before clinical symptom presentation ^12-15^ and other biomarkers are measurable, making them ideal diagnostic biomarker candidates. However, it is increasingly clear that multiplexed panels of Aab biomarkers, are required to provide clinically useful early diagnostic performance^16^, which in turn poses technological challenges in multiplexing classic ELISA measurements, both in terms of sample volumes required and assay costs.

Protein microarrays, in principle, can solve these problems by providing a miniaturised, highly multiplexed, ultra-sensitive ELISA-like assay format and have been used, successfully, to monitor disease activity ^17^ and discover novel autoantibody biomarkers ^18^ in serum. Moreover, previous studies have shown that therapeutic antibody titres measured in DBS are comparable to those measured in serum and plasma ^19,20^, suggesting that the combination of DBS-based sample collection with protein microarray-based autoantibody detection might enable screening of at-risk remote populations for early cancer biomarkers. Here, we describe the development and validation of a robust protein microarray-based method for the quantitation of autoantibody profiles in DBS from patients with chronic pancreatitis (CP) and pancreatic cancer (PC).

## MATERIALS AND METHODS

### Sample Collection and Dried Blood Spots

Inclusion criteria for method development and validation were blood samples from one CP patient prior to resective surgery and 11 randomly selected, late-stage PC patients, respectively (Table 1).

**Table 1.** Demographic and Clinical Characteristics of Patients from which Dried Blood Spots were Extracted.

|  |  |  |
| --- | --- | --- |
| Variable | Disease cohort |  |
|  | Chronic pancreatitis<br>(n=1) | Pancreatic cancer<br>(n=11) |
| Age (y) | 49.0 ± 5.00 | 61.82 ± 11.37 |
| *Sex (n)* |  |  |
| Male | 1 | 4 |
| Female | - | 7 |
| *Race (n)* |  |  |
| Black | 1 | 4 |
| Coloured | - | 5 |
| Other | - | 2 |

This study was approved by the University of CapeTown Human Research Ethics Committee (HREC 559-2018). Written informed consent was obtained from individuals for which study samples were derived.

### In-house DBS cards

Here, 50 µL of whole blood from all 12 patients was pipetted unto Whatman™ filter paper 1 (150 mm; Cat #1001150) and allowed to dry for 1 h, at room temperature (23 □, RT). The filter paper was then placed in resealable plastic bags with a drierite desiccant (#737828-454G, Sigma-Aldrich) and stored in the dark, until ready for further use.

### Commercial DBS cards

ArrayIT blood cards were used to compare and validate the results from our low cost, home-made cards. Following collection, 50 µL of whole blood from the CP patient was pipetted unto ArrayIT dried blood cards (#ABC, ArrayIT corporation) and allowed to separate into red blood cell components and serum for 5 min. The blood cards were then left to dry at RT for 5 min, according to the manufacturers’ instructions, placed in the anti-static shipping envelopes provided, and stored in the dark, until ready for further use.

### Serum control

Following DBS collection, whole blood was centrifuged at 1300 x g for 13 mins, and serum was isolated and stored at -80 □until ready for further use.

### Sample extraction

#### In-house Whatman™ DBS cards

Based on previous DBS methods and the requirements for microarray assays, a method of serum extraction from DBS was developed. On days 1, 5, 10, and 15, a 5mm disc was excised from blood spots on the Whatman™ filter paper using a disposable punch (#MT3336, Integra Miltex). The filter discs were handled with fine-tipped forceps to prevent contamination. Each disc was soaked in 250 µL Phosphate Buffered Saline containing 0.1% Tween 20 (PBST) in a 24-well plate and incubated at RT on a shaker for 60 min × 100 rpm. Subsequently, the eluent was placed in 1.5 mL microfuge tubes and centrifuged at 14 000 g × 10 min to sediment cell debris, after which each supernatant was transferred to a clean tube.

#### Commercial ArrayIT DBS cards

To validate our in-house Whatman™ filter paper method, we extracted serum from ArrayIT blood cards based on the manufacturers’ instructions on days 1, 5, 10, and 15. A 5mm disc was excised from the serum portion of the blood card, presumed to contain antibodies, antigens, and other serum proteins. The disc was wet with 10 µL of PBST, placed in 1.5 mL microfuge tubes, and allowed to re-hydrate for 30 mins at RT. Subsequently, the disc was centrifuged at 14 000 g × 1 min to elute the serum into the collection tube. The disc was then re-wet with 10 µL of PBST and re-centrifuged to elute any remaining bound proteins. The eluents were combined, bringing the final volume to 20 µL.

### Microarray Analysis

To determine the optimal dilution factor of the DBS eluents from each card for the assays, a microarray assay including 5 control spots of known concentrations was performed (Figure S2). We found that, similar to the serum control, a 1:800 dilution of ArrayIT eluent produced an optimal intensity of the control protein whereas a 1:100 dilution of Whatman™ eluent was optimal.

#### Method Development

For the microarray assays, Sengenics IMMUNOME™ arrays pre-printed with 1622 proteins on each array were used. Prior to the assays, the slides were removed from their storage solution and washed 3 × 5 min in PBST and 1 × 5 min in ddH_2_O. The serum and eluents from each method were diluted based on the dot blot assay and slides were incubated with sample on an orbital shaker at 100 rpm for 60 min in a light-protected slide processing dish to prevent photobleaching. Subsequently, slides were washed 3 × 5 min and rinsed 1 × 5 min in PBST and ddH_2_0, respectively. Slides were then incubated in 20 µg/ml of detection antibody (Alexafluor 647 coupled goat anti-human IgG (H + L); Cat#A21445, ThermoFischer Scientific) for 30 min on an orbital shaker at 100 rpm for 30 min. Again, slides were washed in PBST and ddH_2_0, respectively, and then dried via centrifugation at 1300 RCF × 3 min at RT. Dried slides were scanned according to pre-set parameters and saved as TIFF files, which were used for data extraction downstream.

#### Validation

The assay process was replicated on 11 PC patients using a custom-made chip including 3 proteins that had previously been identified (unpublished data) to be upregulated in PC patients (MAGEA5, MART.1, NY.CO.45) and a total anti-human IgG control.

### Statistical Analysis

The microarray image data was extracted using Mapix (v 8.5.0) and the Sengenics IMMUNOME™ gal file, and median foreground and background intensities were read into RStudio (v.4.0) for pre-processing using the ProMAP single channel microarray analysis pipeline script (developed by MOM). Briefly, non-specific binding, array data that was not significantly different to surrounding background (defined as spot intensities <2SD of the median background) were filtered out. Data was then *normexp* background corrected and cyclic loess normalized prior to downstream analysis. A one-way analysis of variance (ANOVA) was used to compare the average log expression intensity of proteins from the three sample collection methods (serum, ArrayIT, and Whatman™ DBS). Subsequently, a limma linear model using calculated array weights, was fit to the normalized microarray data to fully model the systematic part of the data and determine variability between the groups using the limma package in R ^21^. To determine variability in the data based on the comparisons of interest, we extracted contrasts matrices. In this way, we were able to determine if there were differences between: i) eluents from ArrayIT and Whatman™ DBS methods, and ii) eluents from the two DBS methods on Day 1, 5, 10, and 15. Subsequently, an empirical Bayes method was used to moderate the standard errors of the estimated log-fold changes.

Finding no differences between methods or across days we then compared Aab profiles of our Whatman™ method over 15 d. To determine variability in Aab profiles of the 11 PC patients over 15 d for validation of our findings, a limma time course analysis was run. The probability level of p < 0.01 was determined as differentially expressed. All statistical analyses were conducted using R (version 3.6.0) and all result images were created using the ggplot2 package in R ^22^. Power analysis for an effect size = 0.8, yielded a power = 0.805.

## RESULTS

### Comparison of Serum control to Dried blood card methods

Median log expression intensity of proteins from the CP serum control (9.67 ± 0.745) was slightly higher than that of the corresponding ArrayIT (9.60 ± 0.834) and Whatman™ (9.63 ± 0.675) eluents. However, there was no significant difference between the average log expression of the serum control versus ArrayIT (t=-2.820, and p = 0.013) and Whatman™ eluents (t = -1.421, p = 0.330) (F=3.977, p > 0.01; Figure 2a). Furthermore, the MA plot of the protein profiles from all three samples on day 1 showed normalized log fold change values (M) close to zero (Figure 2b). A density plot of the serum control and two DBS methods showed a shift of the peak density to lower average log-expression values for the two DBS methods (Figure 2c), implying that the DBS samples had lower non-specific background signals.

**Figure 2.**
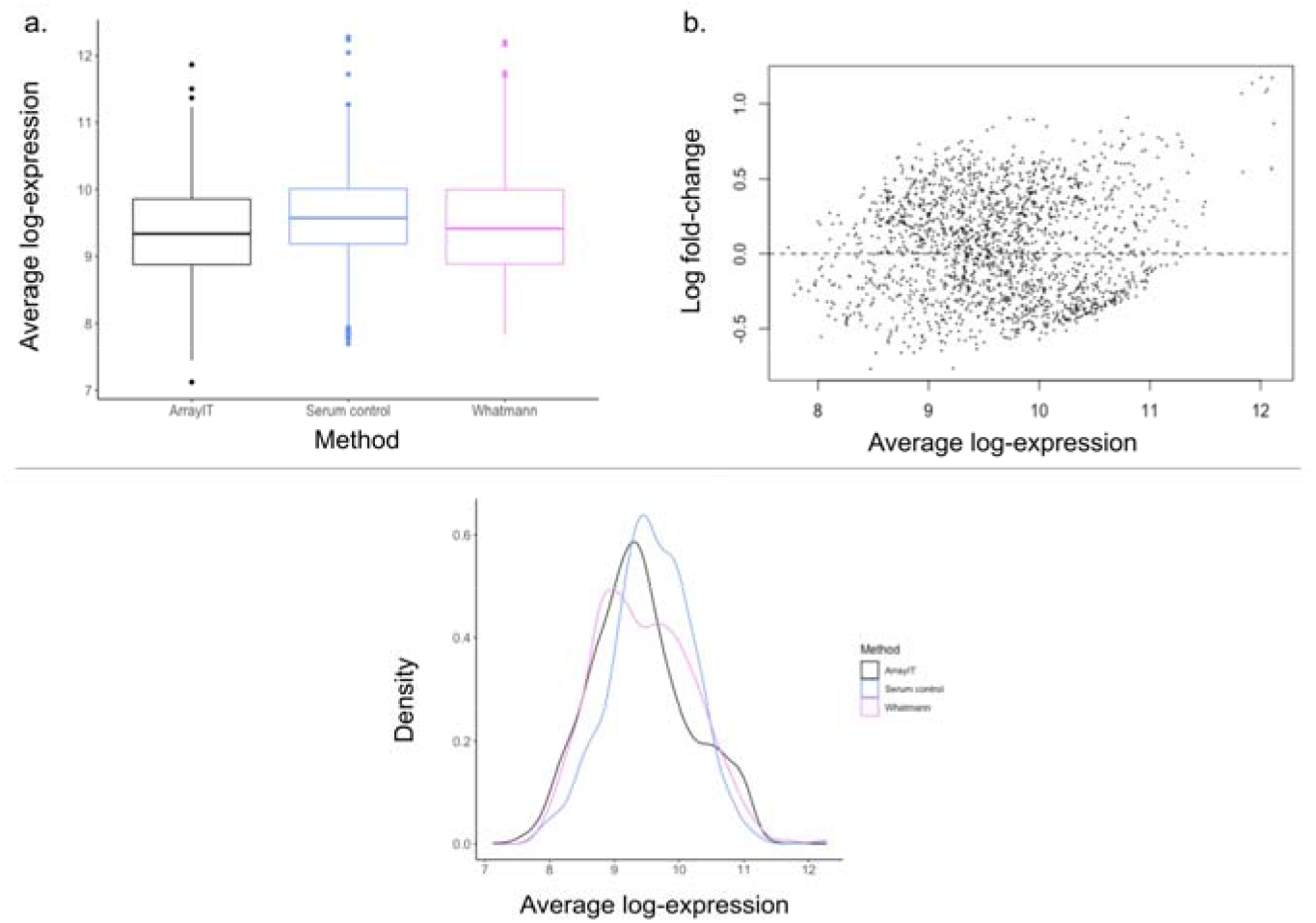
Comparison of average log protein expression from chronic pancreatitis serum control, ArrayIT and Whatman™ eluent samples assayed on the immunome array. a.) Boxplot comparison median average log intensities found in serum sample and ArrayIT and Whatman™ eluent samples b.) MA plot obtained comparing the three extraction methods. M represents the expression intensity of serum versus the average of the two other samples c.) Smoothed empirical densities for average protein log intensities in serum control and DBS arrays.

### Comparison of protein expression of ArrayIT and Whatman™ eluent arrays and over time

There was no significant difference between immunome protein expression of eluents from the ArrayIT and Whatman™ arrays (adj. P > 0.248) over all time points measured (Fig 3a and b). Concurrently, MA plot showed M values close to zero, but slightly higher than the comparison of serum control to both DBS eluents (Figure 3b). Furthermore, we found that there was a slight but non-significant decrease in average log expression over time (adj. P > 0.721) (Fig 3c). Furthermore, there was a slight decline in the abundance of protein expression over time (Figure 3d).

**Figure 3.**
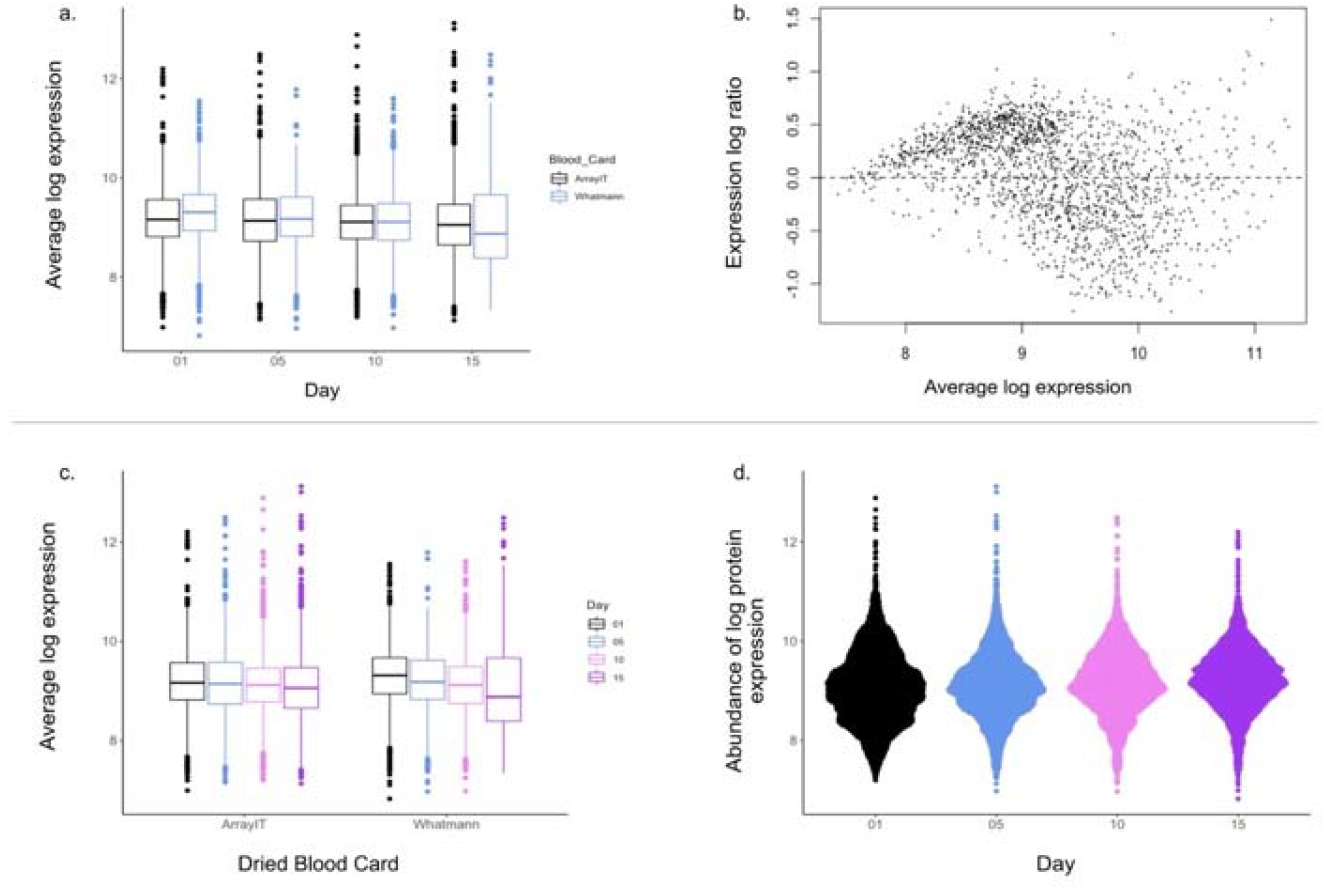
Comparison of average log protein expression of the two dried blood spot eluents from CP patient assayed on the immunome arrays, over time. a.) Boxplot comparing average log expression of ArrayIT and Whatman™ DBS protein arrays b. MA plot obtained comparing the two DBS methods c.) Boxplot comparing protein expression over time in both DBS array types d.) Bee swarm plot visualizing distribution of relative maintained signal for the 1622 proteins analysed compared over 15 d.

### Validation of Whatman™ DBS Method on Custom Microarray

#### Multiple Linear Regression

We ran a multiple linear regression analysis to determine if any factors other than time had an effect on protein expression intensity in the 11 PC patients assayed on the custom DBS array We found no effects of age, race, or gender on protein intensity (F = 0.962, p = 0.493).

#### Time Course Analysis

A limma time course analysis was run to compare protein expression intensities of the 3 proteins and IgG controls assayed over 15 d in 11 PC patients. We found no proteins differentially expressed in any of the time comparisons (Fig 4a). Furthermore, we found no significant difference in expression intensity of any of the proteins over 15 d (p < 0.01, Fig 4b). However, as previously observed we found a reduction, albeit non-significant, in protein abundance over time (Fig 4c).

**Figure 4:**
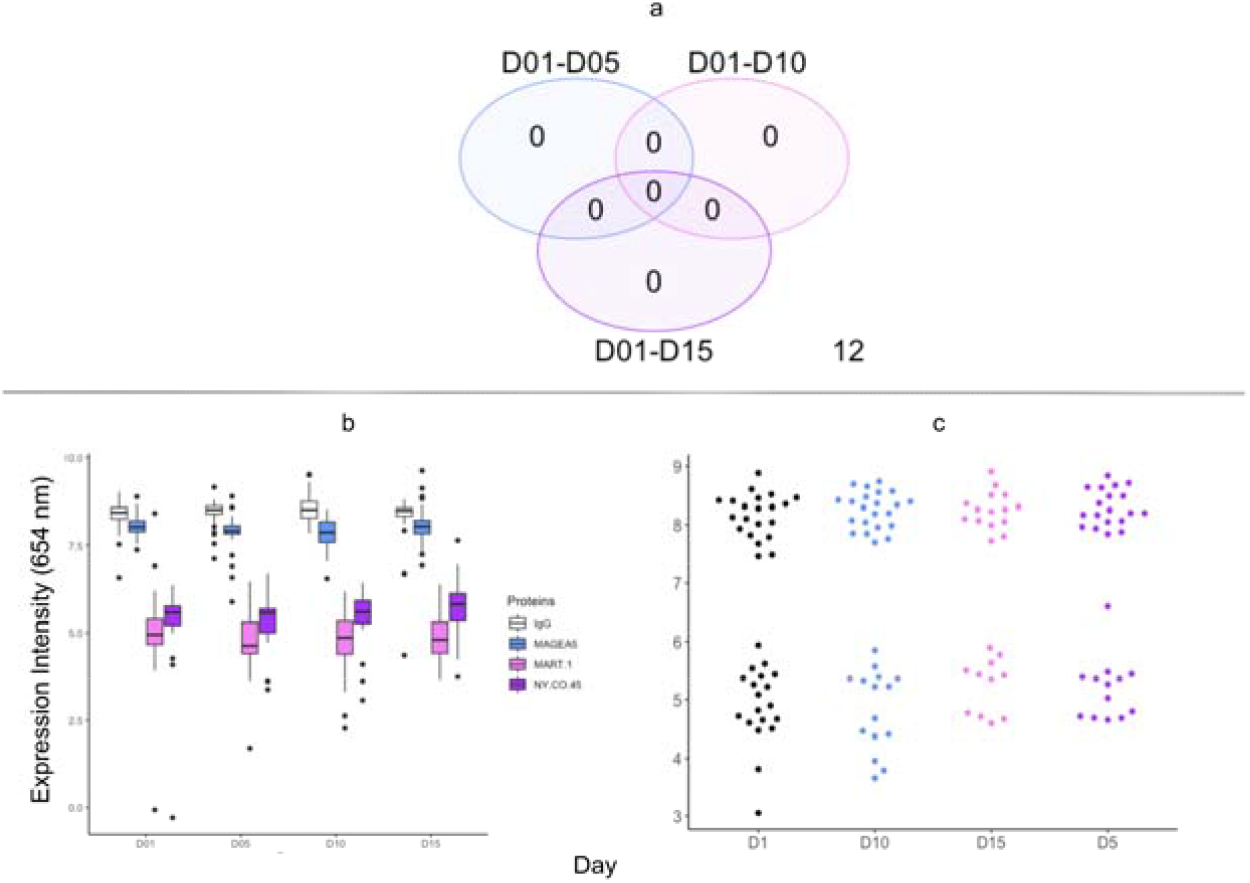
Comparison of protein intensity expression and abundance from day 0 – 15 in 11 pancreatic cancer patients assayed on the custom DBS array. a.) Venn diagram showing number of proteins significantly different in each of the comparisons of days eluted, b.) Boxplot comparing expression intensity of each of the 4 proteins over 15 d, c.) Bee swarm plot visualizing distribution of relative maintained signal for the 4 proteins analysed compared over 15 d

## DISCUSSION

Cancer is an enormous global burden that is predicted to increase with the growth and ageing of populations ^9,23^. Unfortunately, economically disadvantaged countries face a disproportionally high burden of infection-related cancers compared to their developed counterparts. Fortunately, the cancer burden can be largely mitigated by early disease detection ^24^. Serological testing plays an important role in early screening and diagnosis in several disease areas and the identification of more effective biomarkers for early detection. Serum autoantibody profiles can indicate the presence of diseased cells months to years prior to symptom presentation ^12-15^. This is especially useful in rare malignancies, such as PC, known to clinically present at an advanced stage in most patients. Large serosurveys may be key to discovering early detection tools, especially in populations with large rural, remote communities, where access to the associated technologies remains limiting.

This study was performed to quantitatively compare results derived from serum obtained from venous blood collection under routine conditions and blood dried on two different DBS cards, commercial (ArrayIT) and homemade (Whatman™ no 1). Several studies have shown a correlation with data from venous blood samples and blood dried on Whatman™ 903 substrates ^25^. However, Whatman™ no 1 differs from 903 in that the latter is made specifically for protein retention, but the former is more easily accessible largely due to cost and usefulness for various other laboratory experiments. Our use of the latter was influenced by the need for a more costeffective method of blood collection for primarily low-income communities. We developed a method of extracting eluent sample from blood cards made using Whatman™ no 1, eliminating the need for phlebotomies and reducing the sample collection burden on stressed healthcare systems, especially in LMICs.

The results presented here show that eluent extracted from these blood cards up to 15 d following blood collection yielded results comparable to those from serum isolated from whole blood following venous blood collection. In addition, Whatman™ eluents yielded profiles comparable to those of ArrayIT blood cards, thus representing a low-cost alternative to currently available commercial blood cards which typically cost ∼USD10/sample. Notably, protein expression intensities from the serum control sample remained higher, albeit non-significantly, compared to DBS eluents, but simultaneously, the DBS samples appeared to present with a lower background, perhaps due to permanent absorption of larger macromolecules and complement factors on to the membranes.

The dried blood spot modality has the potential for wide scale screening of diseases such as cancers and autoimmune, infectious, and cardiovascular diseases where this is necessary to drastically reduce fatalities. The widespread use of DBS in the past has been impeded by small sample volumes and low target analyte concentrations requiring a sensitive and specific assay for detection and quantification. The use of protein microarray methods for DBS eluent testing has the unique advantage of requiring minimal volumes (1 µl) per assay, without compromising the ultrasensitivity (pg/ml detection range) of the resultant assays ^26^. Thus, DBS samples can be used for multiple testing or to support further analyses. Future studies will investigate the feasibility of a self-collection kit for at-home finger-prick DBS collection using the method developed here to elute and test samples that show comparable analytical performance to venipuncture-derived blood samples.

Overall, this study could pave the way to large scale serosurveys in remote populations by utilizing simple, low-cost DBS sample collection, combined with RT courier shipments to a centralized testing laboratory and miniaturized, protein microarray-based quantitative, multiplexed biomarker detection, thereby increasing the effectiveness of screening campaigns and early diagnosis, and the accuracy of global cancer registries.

## Data Availability

All data produced in this study are available upon reasonable request to the corresponding author.

## ASSOCIATED CONTENT

## AUTHOR INFORMATION

## Author Contributions

**Conceptualization**: JMB, MOM; **Methodology, Validation, Formal analysis, and Investigation**: MOM; **Resources**: MOM, JMB, EJ, UK; **Data curation**: MOM, TR, HA, JG, UK, MB, BA, EJ; **Writing – Original Draft**: MOM; **Writing – Review & Editing**: MOM, JMB, EJ; **Visualization**: MOM; **Supervision**: JMB, EJ; **Project administration**: MOM; **Funding acquisition**: JMB, EJ. The manuscript was written through contributions of all authors. All authors have given approval to the final version of the manuscript.

## ACKNOWLEDGMENTS

We acknowledge all the patients who gave selflessly for the completion of this study. I would also like to thank Dr. Andrew Nel for allowing me to bounce ideas off him during the study. This study was supported by a grant from the Andrea Fine Foundation to JMB and EJ and a National Research Foundation (NRF) South African Research Chair (SARChI) grant [grant number 64760] to JMB. MOM was supported by bursaries from the NRF and Andrea Fine Foundation and MB was supported by a bursary from the Andrea Fine Foundation.

