## Supplemental Figure 1 for "Reaching the Remote: Dried blood spot analysis for disease diagnosis on a protein microarray platform"

Metoboroghene O. Mowoe

PhD Candidate – Chemical and Systems Biology

Institute of Infectious Diseases and Molecular Medicine, Faculty of Health Sciences, University of CapeTown, CapeTown, SouthAfrica

Jonathan M. Blackburn

Deputy Director, Institute of Infectious Disease & Molecular Medicine

Head, Division of Chemical & Systems Biology, Department of Integrative Biomedical Sciences

South African Research Chair in Applied Proteomics & Chemical Biology, University of Cape Town

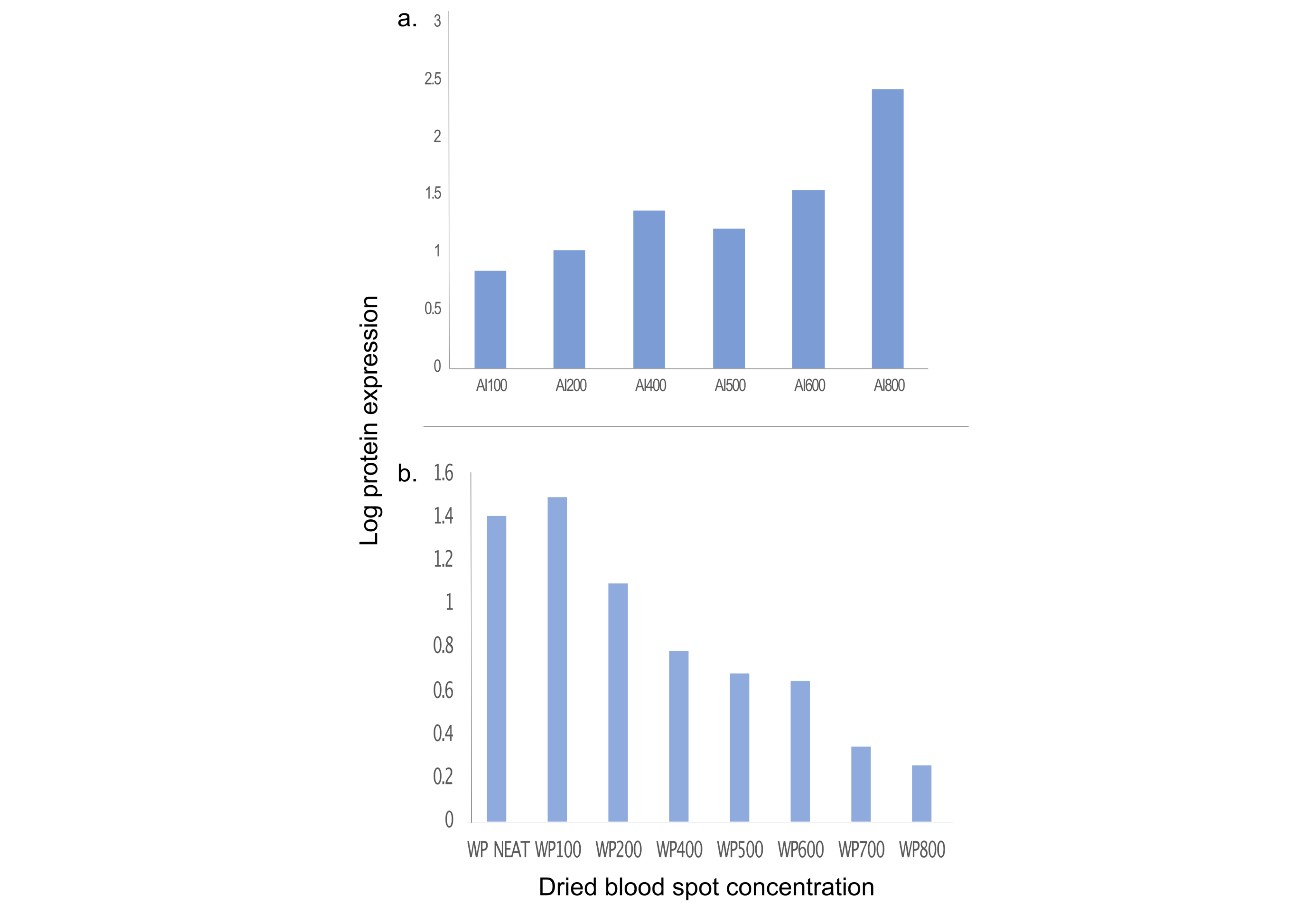

**Figure S1.** Protein expression to determine optimal dilution for a.) ArrayIT and b.) Whatmann dried blood spot eluents for subsequent microarray assays.

*Ai(n) – ArrayIT dried blood card (dilution factor); W ()– Whatmann filter paper dried blood card (dilution factor).
